# Study Report from the Pragmatic Assessment of the NuvoAir Clinical Service in the Management of Patients with Chronic Obstructive Pulmonary Disease

**DOI:** 10.1101/2025.11.11.25339788

**Authors:** Eric Harker, Jeffrey J. Van Wormer, Dandi Qiao

## Abstract

**Background:** COPD is one of the leading causes of death, disability and avoidable hospitalization in the US and abroad, yet proactive management of COPD has historically been limited. This study evaluates NuvoAir Home Service (NAHS), a novel intervention that integrates telemedicine, remote monitoring, and chronic-care management to deliver proactive care to patients with COPD.

**Methods:** We present findings from a six-month pragmatic program evaluation. Patients with COPD enrolled in NAHS were compared with propensity-matched controls using a two-period difference-in-difference (DID) design.

**Results:** 280 NAHS participants compared to 840 propensity matched controls showed a reduction of 47% in post-to-pre risk ratio (RR) for COPD-related hospitalization (p = 0.06), 55% in average days of hospital stay (p = 0.02), 53% in hospital readmissions (p = 0.32). There was no significant change in ED visits (p=0.83), and in outpatient office visits (p=0.89). All-cause utilization moved in similar directions, but none of those changes were statistically significant. COPD-related total medical cost and hospitalization cost from medical claims were also significantly reduced with a 28% reduction (p-value = 0.05) and a 53% reduction (p-value = 0.02) in post-to-pre cost ratio. No significant changes were observed in all-cause hospitalization, all-cause and COPD-related ED, outpatient, or pharmacy costs.

**Conclusion:** These data indicate that NuvoAir Home Service had a clinically positive impact on COPD-related hospital utilization and costs without significantly increasing ED or outpatient use, though not all results met statistical significance.

## 1. Introduction

The US healthcare system struggles to adequately address the burden of Chronic Obstructive Pulmonary Disease (COPD), and the challenges in managing COPD are growing, while resources are diminishing. COPD is the third leading cause of death in the world behind only cardiovascular diseases and cancer.[1] COPD is a leading cause of disability, and according to the World Health Organization is the third leading cause of healthy life lost to combined disability and death, as measured by Disability Adjusted Life Years.[2] Yet management of COPD can be suboptimal, with both primary care and specialty physicians often failing to follow clinical guidelines.[3] In a study of 238,158 newly diagnosed patients with COPD, 86.2% of patients were initially untreated, and most (63.8%) remained untreated at the end of a 4-year follow-up period, despite 33% having a moderate to severe exacerbation.[4] Access to high quality care in rural areas of the US is particularly poor, with rural residents more likely to forgo doctor visits due to cost, more likely to be uninsured, and only 35% of rural residents living within 10 miles of a pulmonologist.[5,6]

COPD is extremely expensive, estimated to account for $50 billion in healthcare spending in 2020, and is one of the leading causes of potentially avoidable hospitalization. Hospitalizations, which account for >60-70% of all COPD-related medical costs,[6] are often triggered by COPD exacerbations. But those exacerbations are also associated with subsequent cardiovascular complications including acute coronary syndrome, congestive heart failure, and stroke.[7] 80% of patients with COPD suffer from comorbid conditions, including congestive heart failure, coronary artery disease, peripheral vascular disease, stroke, hypertension, and diabetes.[8]

Caring for patients with COPD requires effectively managing comorbid cardiovascular and related conditions, thereby requiring *cardiopulmonary* solutions. In recent years, comprehensive remote care management has gained attention as a potential tool to improve outcomes, reduce hospital admissions and lower the cost of care for COPD and related comorbid conditions.[9] Accordingly, CMS has implemented reimbursement models for remote care management, including Remote Physiologic Monitoring (RPM) and Chronic Care Management (CCM). However, some studies have shown that proactive care management of COPD can lead to *worsening* outcomes, including increased hospitalization, ED visits and even mortality. [10,11] There is some evidence that outcomes may be *worse* if self-care and monitoring is promoted without also providing enhanced timely *access to care* for signs of exacerbation. For this reason, the NuvoAir solution required that exacerbations not responding to standardized COPD Action Plans either receive same day care by a patient’s existing care team, or same day telemedicine care by NuvoAir physicians and nurse practitioners.

## 2. Methods

### 2.1. Design

This is a pragmatic evaluation of the NAHS program for patients with COPD living in north-central Wisconsin. Specifically, all participants were members of Security Health Plan (SHP) of Wisconsin, Inc., an affiliate of the Marshfield Clinic Health System. A cohort evaluation was used, including NAHS enrollees from July 2023 to October 2024 vs. a propensity score matched comparison group of similar individuals not enrolled in NAHS. The original study was designed to assess the intervention’s effects over 12 months; however, due to unforeseen business circumstances, the available sample size only supports a 6-month evaluation in the final study.

### 2.2. Participants

All study participants had evidence of COPD in the health insurer’s database, per medical diagnostic codes. Because this was a pragmatic evaluation of real-world COPD care, and because a significant proportion of patients diagnosed with COPD do not have documented spirometry confirmation of COPD diagnosis in the US,[12] spirometry confirmation of COPD diagnosis was not required. However, remote, in-home spirometry testing was offered to all participants, delivered to their homes at no cost. Participants unable to perform remote spirometry due to poor tech literacy or lack of access to the internet were encouraged to receive in-office spirometry from their usual care providers. Spirometry results were used in this program to improve the diagnosis and treatment of all patients, including potential de-escalation and re-evaluation of patients found to potentially be mis-diagnosed with COPD based on spirometry results. We followed an intention to treat approach and did not exclude patients without spirometry confirmed COPD, in part because spirometry confirmation was not possible for all patients, particularly those in the control group, for whom we only had access to de-identified claims data.

All participants had reasonable access to usual medical care given their insured status. Study participants were recruited via outbound email and text enabled IVR calls, based on COPD diagnoses in their claims data. The original study design sought only high-risk COPD patients, defined by those with a COPD related hospitalization in the past 12 months, but to reach recruitment goals, the population had to be expanded to all patients with COPD related claims. High risk patients with claims received additional, focused outbound recruiting messages compared to others. Beyond this, there was no influence on recruitment by study personnel, and no patients were referred to the program by their routine clinical teams. Those in the NAHS group received their routine clinical care from their usual healthcare providers, with the addition of the NuvoAir Home Service.

In contrast, those in the non-NAHS group received routine care from their usual healthcare providers, and only anonymized claims data collected by their health insurer was subject to analysis. NuvoAir Home and standard care groups were propensity score matched based on sex, days from last inpatient event to enrollment, and disease severity (12-month pre-enrollment: number of outpatient visits, ED, number of hospitalizations - all cause and COPD-specific). Controls were matched in parallel and followed over the same period. Historical controls, including pre-post study design, were not elected for two reasons: First, confounding effects of evolving COVID-19 infection prevalence would complicate historical comparisons, and second, patients with COPD tend to progress over time with an accelerating pattern of exacerbations and complications, making a historical comparison problematic.

Both groups were selected from the same rural geographic areas in north-central Wisconsin, surrounding Marshfield Wisconsin, a town of under 20,000 people with the nearest metropolitan area over 80 miles away. Participants and controls were predominantly from elderly, rural population, often with limited access to and familiarity with digital health technology. As this was a real-world evaluation, not focused only on those with access to and familiarity with technology, participants were not excluded based on technical abilities, and instead were offered low tech options, including care by telephone only, when needed. This study was a quality improvement program evaluation and determined to be exempt by the Marshfield Clinic Health System Institutional Review Board.

### 2.3. NuvoAir Home Service NAHS

NuvoAir Home Service (NAHS) is a technology-driven, team-based remote care intervention designed to support patients with heart and lung conditions through at-home monitoring, remote chronic care management, and specialty pulmonary care via telemedicine. The purpose of this study was to evaluate the early outcomes from a pilot program testing NAHS. As hospital admissions are the primary driver of COPD-related costs, this evaluation has a particular focus on early reductions of avoidable hospital admissions as the primary outcome.

The NuvoAir Home Service is designed to incorporate key components of evidence-based care for COPD, including maintenance bronchodilators with or without inhaled steroids for patients with GOLD group B and E COPD, multidisciplinary team-based care to support patient self-management, and remote monitoring with telehealth.[13,14] The NAHS remote monitoring program includes daily heart rate and pulse oximetry testing and weekly home-based spirometry testing, which has been shown to detect COPD exacerbations up to 2 weeks prior to reported symptoms.[15] Additionally, patients are asked to complete a digital version of the COPD Foundation’s My COPD Action Plan daily based on patient reported symptoms of dyspnea, cough, functional decline and others. The My COPD Action Plan is a key component of supported self-management programs, which are recommended by international guidelines, including GOLD.[16] These biomarkers and patient reported symptoms and outcomes were collected via smart phone and tablet-based apps, or by text for those not able to use app-based technology. Patient-supplied data was collected by the NuvoAir Home Portal, monitored using algorithms and by the coaches, and signals of respiratory exacerbation or other clinical decline, such as decreases in FEV1, pulse oximetry, or COPD Action Plan zone were reported to the NuvoAir team for appropriate outreach, assessment, triage, and management by COPD Action Plan protocol.

The NuvoAir Home Service also includes remote interaction with a dedicated NuvoAir care coordinator who provides support and education, including monthly assessment and educational sessions where COPD Foundation educational materials were presented and reviewed, including topics such as breathing techniques, coping with illness, preventing exacerbations, COPD medication use, preventing pneumonia, and more.[17] Daily monitoring of symptoms, heart rate, and pulse oximetry as well as weekly spirometry was encouraged and regular communication was provided via telephone, synchronous video visit, text, and chat, depending on the preferences and abilities of individual participants. A NuvoAir Registered Nurse provides medical advice and triage for patients experiencing new or worsening symptoms or signals of decline detected by the NuvoAir digital platform. Finally, a NuvoAir physician or nurse practitioner provides consultation and urgent telemedicine evaluation and treatment for those patients without access to same-day primary or specialty care for suspected COPD exacerbations. Patients with clinical evidence of COPD exacerbation were referred to same day care by their existing care teams, or if needed, treated by NuvoAir standardized protocol, consisting of breathing exercises and activity modification, optimization of rescue inhalers, and if needed, short courses of prednisone and antibiotics.

## 3. Outcomes

This report focuses on participants who have completed at least six months of participation in the study. The primary end points were all-cause and COPD-related rates of hospitalizations and Emergency Department (ED) visits. Secondary end points included the rate of hospital readmission in 30 days, average days of hospital stay, the rate of outpatient office visits. Exploratory end points include utilizations costs, pharmacy costs, and medication adherence to COPD maintenance medications.

All clinical care utilization data were computed based on the de-identified medical and pharmacy claims provided by SHP. Hospitalizations are referring to inpatient stays that were at least 30 days apart. If an ED visit was followed by an inpatient stay, the event was counted toward a hospitalization event. Average days of hospital stay is the number of days the patient was hospitalized in a single inpatient stay. The total number of outpatient office visits were counted along with the number of outpatient office visits with a COPD exacerbation diagnosis or followed by a short-term prescribed course of antibiotics or corticosteroids (namely acute COPD outpatient office visit). The total pharmacy cost and COPD-related pharmacy cost were obtained using the pharmacy claim for the same patients, along with a curated list of pulmonary medications usually prescribed to COPD patients.

To identify if the intervention has any effect on patients’ medication adherence to COPD maintenance medications, two metrics (Medication Possession Ratio (MPR) and Proportions of Days Covered (PDC)) were computed using the pharmacy claims provided. A patient is defined as medication adherent in MPR (PDC) if maximum MPR (PDC) > 0.8 for the patient among all COPD maintenance medications the patient filled during the period. The percentage of patients who were medication adherent were compared before and after the intervention.

## 4. Analyses

Patients in the intervention group were enrolled and engaged patients who have provided biomarker data or have more than two verbal communications with the NuvoAir clinical team. The index date was defined as the latest date among the patient’s enrollment date, first communication date, and biomarker synchronization date. Only patients whose index date before October 1, 2024, were included as claim data for events before April 1, 2025, needed to be available for 6-month analyses in July 2025. These patients were also required to be eligible for the insurance for the 12 months prior to index date and the 6 months after.

Patients in the standard care group were selected using propensity score matching [18] (PSM) with a ratio of 1:3 from the set of patients who have never enrolled in NuvoAir service provided by SHP. Matching was conducted based on both demographic variables and clinical variables related to disease severity and health care utilizations, and the 6-month study period (to match the seasonal effects in the intervention group). Demographic variables include sex, age, and whether living in a rural or urban area. Clinical variables include: number of hospitalizations, ED visits, and outpatient visits related to all causes, potentially impacted by the service, and those related specifically to COPD in the 12-months prior to the start of the study; number of pulmonary medications and rescue inhaler filled in the 12-months prior to the start of the study; and the number of days from the last day of any hospitalization to the start of the study. A logistic regression with the treatment group flag was fitted based on these matching variables to obtain the propensity score, and a greedy 1-to-3 nearest neighbor algorithm without replacement was used to select the best matching controls with the same index month as the intervention group.

Summary statistics of demographic and baseline clinical variables before and after PSM are presented in mean and standard deviation. For cost variables, mean is the per-member-per-year (PMPY) value, and additional summary statistics such as conditional median and IQR for before and after PSM are presented in the Supplement. To test whether there is any difference in these variables between the intervention group and the controls, Fisher’s exact test was applied to binary variables; t-test test was applied to continuous variables such as age; non-parametric Mann-Whitney U test was applied to count and cost variables. Before matching, the control group refers to all patient-month pairs in which the data for the year starting from the month is available for the control. After matching, the control group refers to the matched controls only.

For all outcomes except the number of readmissions, multivariable difference-in-difference (DID) regression models were applied to compare pre-post changes between the intervention group and control group. [19, 20] Specifically, a generalized linear mixed effects model (GLMM) with a random intercept for each patient was fitted:

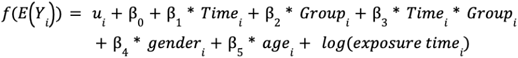

where *Yi* is the outcome of interest; *f* is the link function appropriate for the distribution assumption of the outcome; *u*_*i*_ is a random intercept for each patient following a normal distribution; *Time*_*i*_ is 0 for the pre-12 month period and 1 for the post-engagement period; *Group*_*i*_ is 0 for the control group and 1 for the intervention group. *Exposure time*_*i*_ represents the number of years for the period, 1 for 12-month period and 0.5 for 6-month period.

For count variables with large enough counts, a Negative Binomial or a Poisson distribution (log-link) was applied based on the goodness of fit of the model; for continuous variables with more than 5% of zeros, a Tweedie distribution (log-link) was applied as these variables analyzed are highly skewed; for continuous variables with less than 5% of zeros, a Gamma distribution is applied to the modified cost variable with an offset of 0.01; for binary variables, logistic link was applied. The coefficient for the interaction term captures the average effect of the intervention. These DiD models remove time-invariant confounding through pre-post differences and can improve power while reducing the risk of false-positive findings - provided the parallel trend assumption holds.

For all-cause or COPD-related readmissions, because the number was very low for GLMMs, we compared the incidence rates (events per patient-time) during the 6-month follow-up using an exact two-sample Poisson test. This test provides an exact rate ratio for the intervention versus control group with exact 95% confidence interval and p-value, avoiding the small-sample limitations of GLMMs.

As sensitivity analyses, a different set of models was applied to ensure the results are not highly dependent on the model assumptions. Here a direct comparison between the post 6-month period was conducted using GLMMs with appropriate link functions for different distributions (namely case-control (cc) models here). The matching strata was included as the random effect which preserves the 1-to-3 paired structure created by matching. Covariates such as age, disease severity, number of days from the last day of hospitalization to the start of intervention were included as fixed effects.

In addition, we conducted another sensitivity analysis to obtain bootstrapped effect size estimates and p-values.[18] In each bootstrap, the original intervention group and all controls were sampled with replacement, the propensity score was re-estimated, the same 1:3 matching process was conducted to select the matched controls, and DID model was applied to the matched samples. The bootstrapped effect sizes were obtained using the averaged effect sizes, and p-values were obtained using the distribution of effect sizes across all bootstrap replicates. This method takes into account the uncertainty from both the propensity model estimation and matching process, to ensure the results do not depend on a particular matched sample. 200 bootstraps were run for this analysis.

For all tests, a two-sided significance level of 0.05 was applied to declare statistical significance. Data transformation and analyses were conducted using Python 3.9 and R version 4.3.1 (R package glmmTMB for GLMMs).

### 5. Results

### 5.1. Propensity score matching and baseline characteristics

There were a total of 852 patients participating in NAHS and 6648 patients in the standard care group. With the inclusion criteria in engagement and the availability of claim data for the 12 months before engagement and 6 months after engagement, 280 of the 852 patients were included in the intervention group. Figure 1 shows the flow chart of the inclusion criteria and the number of patients included in the analyses.

**Figure 1.**
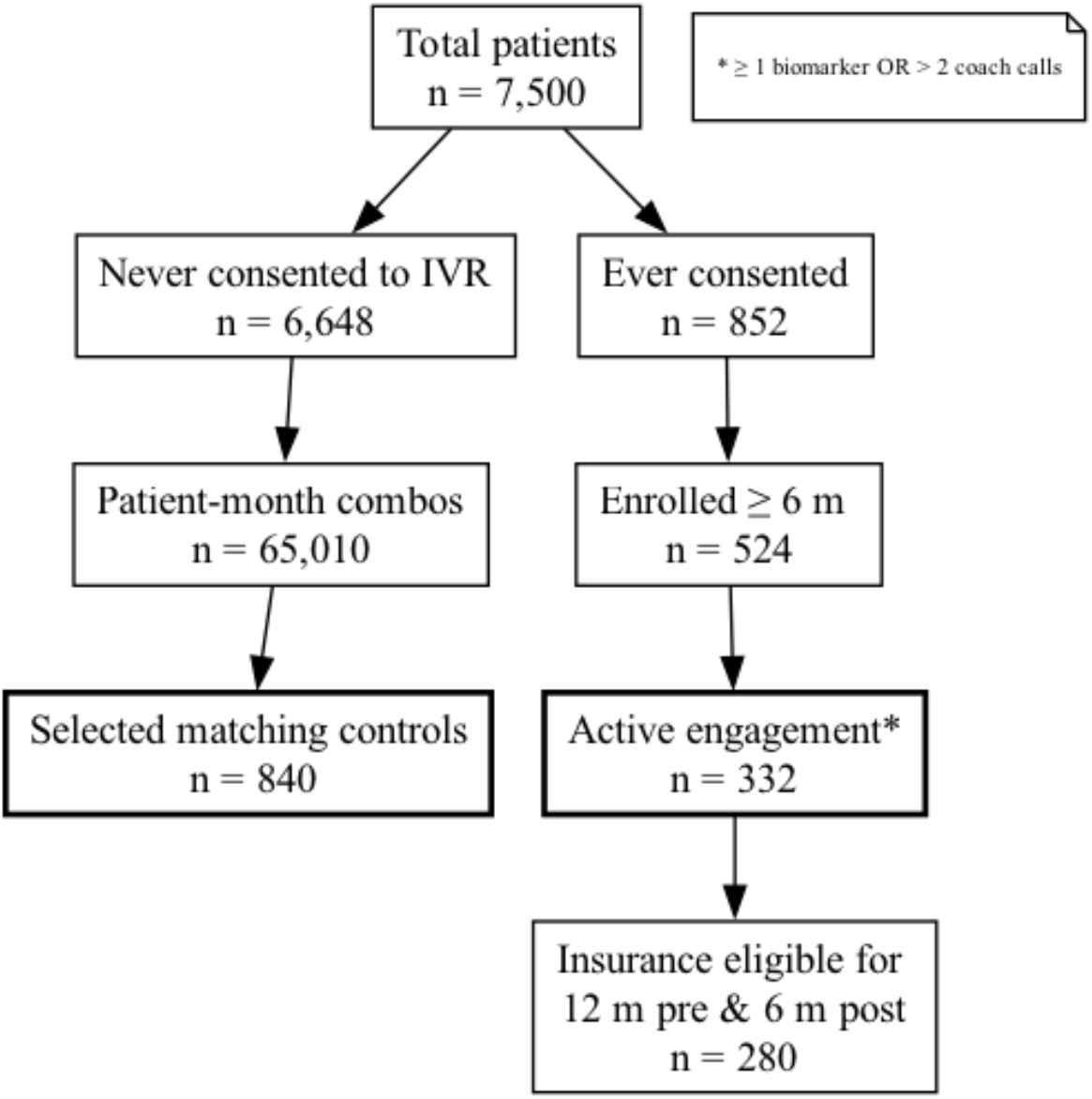
Flow diagram of inclusion criteria for patients in the analyses.

Before any matching, patients in the standard care group required fewer clinical care utilizations and less utilization and pharmacy cost compared to the intervention group (Table 1, Supplemental Table 1). Using PSM, 840 unique patients from the standard care group were selected as the control group. Table 1 shows the comparison in demographic and baseline clinical variables between the intervention group and the control group before and after PSM. There was no significant difference in any of the demographic or baseline variables after matching (Table 1, Supplemental Table 2).

**Table 1.** Demographic and baseline clinical characteristics in the intervention and control groups in the 12 months prior to engagement date, before and after PSM.

|  | Before PSM |  |  | After PSM |  |  |
| --- | --- | --- | --- | --- | --- | --- |
| Characteristic | Intervention | Control | P value | Intervention | Control | P value |
| N | 280 | 65010 |  | 280 | 840 |  |
| Demographic and baseline characteristics |  |  |  |  |  |  |
| Age (Mean ± SD) | 76.82 ± 8.27 | 77.75 ± 8.18 | 0.062 | 76.82 ± 8.27 | 76.32 ± 8.24 | 0.38 |
| Sex Female, n (%) | 137 (48.9) | 32681 (50.3) | 0.6753 | 137 (48.9) | 389 (46.3) | 0.448 |
| Region Rural, n (%) | 192 (68.6) | 42090 (64.7) | 0.1884 | 192 (68.6) | 564 (67.1) | 0.7127 |
| Asthma diagnoses Yes, n (%) | 47 (16.8) | 8634 (13.3) | 0.09317 | 47 (16.8) | 127 (15.1) | 0.5058 |
| CHF diagnoses Yes, n (%) | 89 (31.8) | 19229 (29.6) | 0.4312 | 89 (31.8) | 261 (31.1) | 0.8236 |
| Baseline clinical care utilizations |  |  |  |  |  |  |
| All-cause hospitalizations (Mean ± SD) | 0.28 ± 0.59 | 0.28 ± 0.59 | 0.9 | 0.28 ± 0.59 | 0.29 ± 0.58 | 0.52 |
| All-cause average days hospitalized (Mean ± SD) | 1.18 ± 3.41 | 1.13 ± 3.11 | 8.80E-01 | 1.18 ± 3.41 | 1.04 ± 2.59 | 0.67 |
| All-cause readmissions (Mean ± SD) | 0.04 ± 0.22 | 0.05 ± 0.34 | 5.70E-01 | 0.04 ± 0.22 | 0.04 ± 0.26 | 0.85 |
| All-cause ED (Mean ± SD) | 0.82 ± 1.18 | 0.71 ± 1.32 | 0.012 | 0.82 ± 1.18 | 0.75 ± 1.29 | 0.11 |
| All-cause outpatient office visit (Mean ± SD) | 9.09 ± 5.84 | 7.41 ± 5.94 | 4.20E-09 | 9.09 ± 5.84 | 9.14 ± 6.84 | 0.35 |
| COPD-related hospitalizations (Mean ± SD) | 0.14 ± 0.41 | 0.13 ± 0.39 | 0.55 | 0.14 ± 0.41 | 0.13 ± 0.40 | 0.73 |
| COPD-related average days hospitalized (Mean ± SD) | 0.48 ± 1.43 | 0.66 ± 2.70 | 0.71 | 0.48 ± 1.43 | 0.44 ± 1.43 | 0.52 |
| COPD-related readmissions (Mean ± SD) | 0.01 ± 0.12 | 0.02 ± 0.20 | 0.7 | 0.01 ± 0.12 | 0.01 ± 0.15 | 0.63 |
| COPD-related ED (Mean ± SD) | 0.21 ± 0.50 | 0.13 ± 0.43 | 4.60E-05 | 0.21 ± 0.50 | 0.21 ± 0.60 | 0.46 |
| COPD-related outpatient office visit (Mean ± SD) | 1.06 ± 1.50 | 0.60 ± 1.09 | 3.40E-09 | 1.06 ± 1.50 | 1.04 ± 1.49 | 0.93 |
| Acute COPD outpatient office visit (Mean ± SD) | 0.37 ± 0.98 | 0.25 ± 0.99 | 6.00E-05 | 0.37 ± 0.98 | 0.50 ± 1.41 | 0.25 |
| Rescue inhaler filled Yes, n (%) | 189 (67.5) | 31122 (47.9) | 5.52E-11 | 189 (67.5) | 547 (65.1) | 0.513 |
| COPD-related antibiotics or corticosteroid course (Mean ± SD) | 0.95 ± 1.31 | 0.73 ± 1.36 | 0.00019 | 0.95 ± 1.31 | 1.13 ± 1.66 | 0.26 |
| COPD medication adherent (MPR > 0.8), n (%) | 111 (39.6) | 15878 (24.4) | 2.59E-08 | 111 (39.6) | 307 (36.5) | 0.3547 |
| COPD medication adherence (PDC > 0.8), n (%) | 90 (32.1) | 12214 (18.8) | 9.30E-08 | 90 (32.1) | 246 (29.3) | 0.3672 |
| Baseline cost of clinical care utilizations |  |  |  |  |  |  |
| All-cause medical cost (Mean ± SD) | 15331.48 ± 20044.61 | 16122.35 ± 30943.12 | 0.0011 | 15331.48 ± 20044.61 | 18968.46 ± 33347.24 | 0.68 |
| All-cause hospitalization cost (Mean ± SD) | 5423.00 ± 12744.40 | 5620.02 ± 17238.90 | 0.98 | 5423.00 ± 12744.40 | 5478.60 ± 13762.87 | 0.62 |
| All-cause ED cost (Mean ± SD) | 1053.59 ± 2030.50 | 936.09 ± 2512.37 | 0.015 | 1053.59 ± 2030.50 | 1004.68 ± 2346.93 | 0.15 |
| All-cause office visit cost (Mean ± SD) | 2274.86 ± 2981.09 | 3221.24 ± 14639.33 | 7.20E-08 | 2274.86 ± 2981.09 | 4751.62 ± 22361.20 | 0.79 |
| All-cause pharmacy cost (Mean ± SD) | 10967.93 ± 34747.37 | 5781.39 ± 26302.61 | 1.40E-06 | 10967.93 ± 34747.37 | 7799.49 ± 31644.67 | 0.34 |
| COPD-related medical cost (Mean ± SD) | 3610.45 ± 8845.07 | 3288.24 ± 11882.33 | 1.90E-08 | 3610.45 ± 8845.07 | 3232.72 ± 9256.17 | 0.22 |
| COPD-related hospitalization cost (Mean ± SD) | 2358.92 ± 7721.14 | 2477.29 ± 11165.13 | 0.61 | 2358.92 ± 7721.14 | 2058.59 ± 8280.59 | 0.57 |
| COPD-related ED cost (Mean ± SD) | 288.21 ± 893.45 | 191.14 ± 884.58 | 5.00E-05 | 288.21 ± 893.45 | 254.32 ± 841.59 | 0.42 |
| COPD-related office visit cost (Mean ± SD) | 249.53 ± 389.95 | 187.43 ± 993.04 | 8.60E-09 | 249.53 ± 389.95 | 359.21 ± 1982.87 | 0.92 |
| COPD-related pharmacy cost (Mean ± SD) | 2855.88 ± 4178.68 | 1649.37 ± 3538.24 | 1.70E-09 | 2855.88 ± 4178.68 | 2479.67 ± 4223.69 | 0.33 |

For the 1120 patients included in the analyses, 98% are white and 67.5% live in rural areas in Wisconsin. Most participants were between the ages of 70 and 90, with significant numbers above 90. About 30% of these patients also had a congestive heart failure (CHF) diagnosis in the 12 months before the start of the study as indicated in the medical claim data.

### 5.2. study results

In the 6-month follow-up period, 11.8% and 15.2% of patients had at least one all-cause hospitalization in the intervention group and in the matched control group respectively; 1.8% and 2.7% of patients had at least one all-cause readmission respectively. The event counts and incidence rates are shown in Table 2. Also, a clinical significant reduction in post/pre rate ratios was observed for all-cause average days of hospital stay and COPD-related readmissions. However, these variables did not reach statistical significance (Figure 2, Table 3). The post/pre rate ratio in COPD-related hospitalizations was reduced by 46.8% in the intervention group with a near-significance p-value of 0.060. The post/pre ratio in average days of hospital stay was reduced by 55.4% with a significant p-value of 0.022. These observations are in line with the significant reduction observed in post/pre cost ratios in COPD-related medical cost (28.0% reduction, p-value = 0.047) and COPD-related hospitalization cost (53.1% reduction, p-value = 0.022). There was no significant reduction in all-cause pharmacy cost, COPD-related pharmacy cost, or percentage of patients who are adherent to COPD maintenance medications (Figure 3, Table 3).

**Table 2.** Medical events counts and incidence rate in the 6-month follow-up period.

| Variable | Total number of events |  | Number of patients with events |  | Incidence rate per 1000 patient-year |  |
| --- | --- | --- | --- | --- | --- | --- |
|  | Intervention | Control | Intervention | Control | Intervention | Control |
| All-cause hospitalizations | 41 | 150 | 33 (11.8%) | 128 (15.2%) | 292.86 | 357.14 |
| All-cause readmissions | 6 | 35 | 5 (1.8%) | 23 (2.7%) | 42.86 | 83.33 |
| All-cause ED visits | 126 | 366 | 82 (29.3%) | 234 (27.9%) | 900 | 871.43 |
| All-cause outpatient office visits | 1289 | 3928 | 264 (94.3%) | 778 (92.6%) | 9207.14 | 9352.38 |
| COPD-related hospitalizations | 15 | 79 | 13 (4.6%) | 70 (8.3%) | 107.14 | 188.1 |
| COPD-related readmissions | 3 | 19 | 3 (1.1%) | 13 (1.5%) | 21.43 | 45.24 |
| COPD-related ED | 31 | 89 | 22 (7.9%) | 72 (8.6%) | 221.43 | 211.9 |
| COPD-related outpatient office visit | 140 | 405 | 90 (32.1%) | 257 (30.6%) | 1000 | 964.29 |
| Acute COPD outpatient office visit | 50 | 192 | 35 (12.5%) | 112 (13.3%) | 357.14 | 457.14 |

**Table 3.**
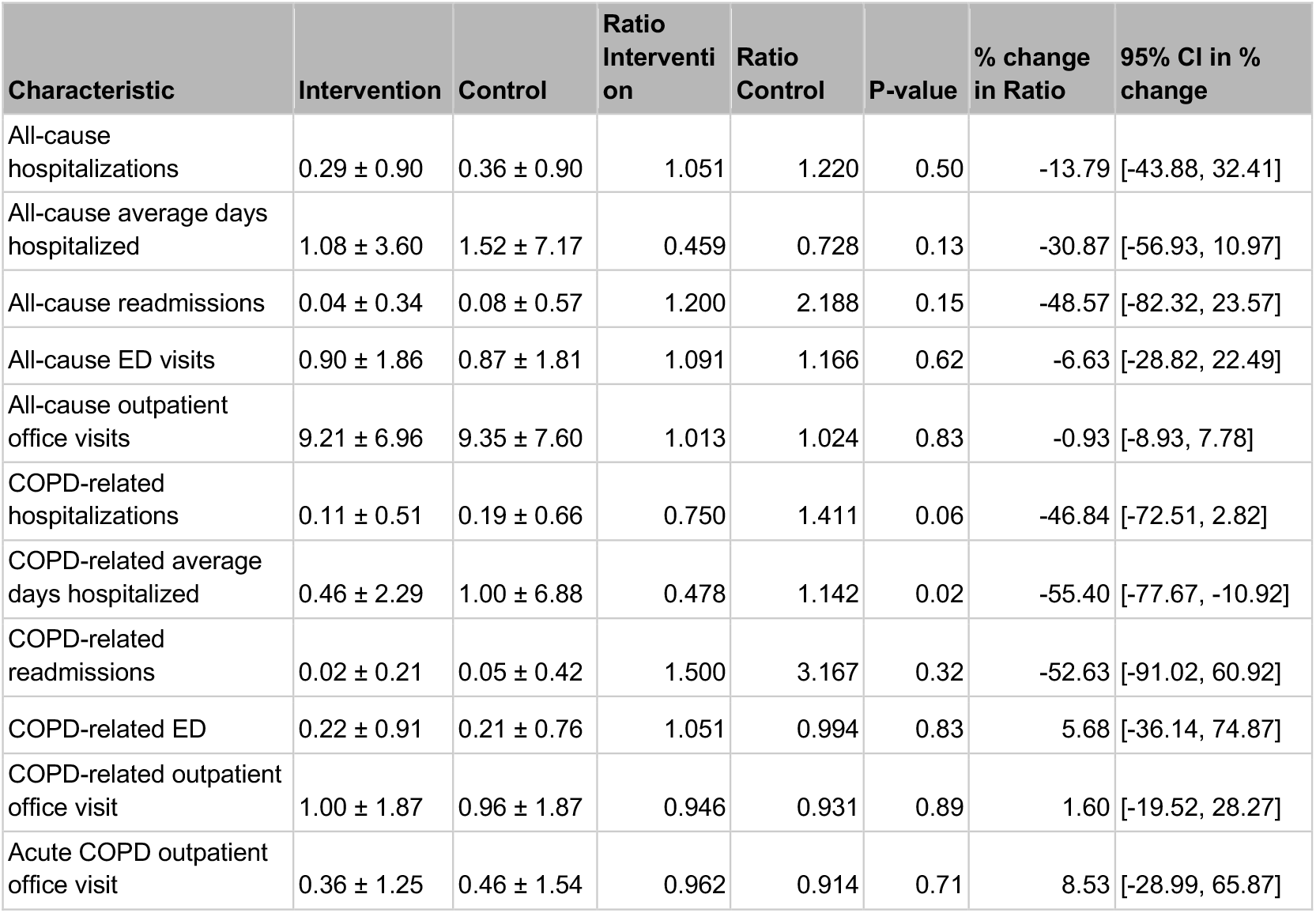
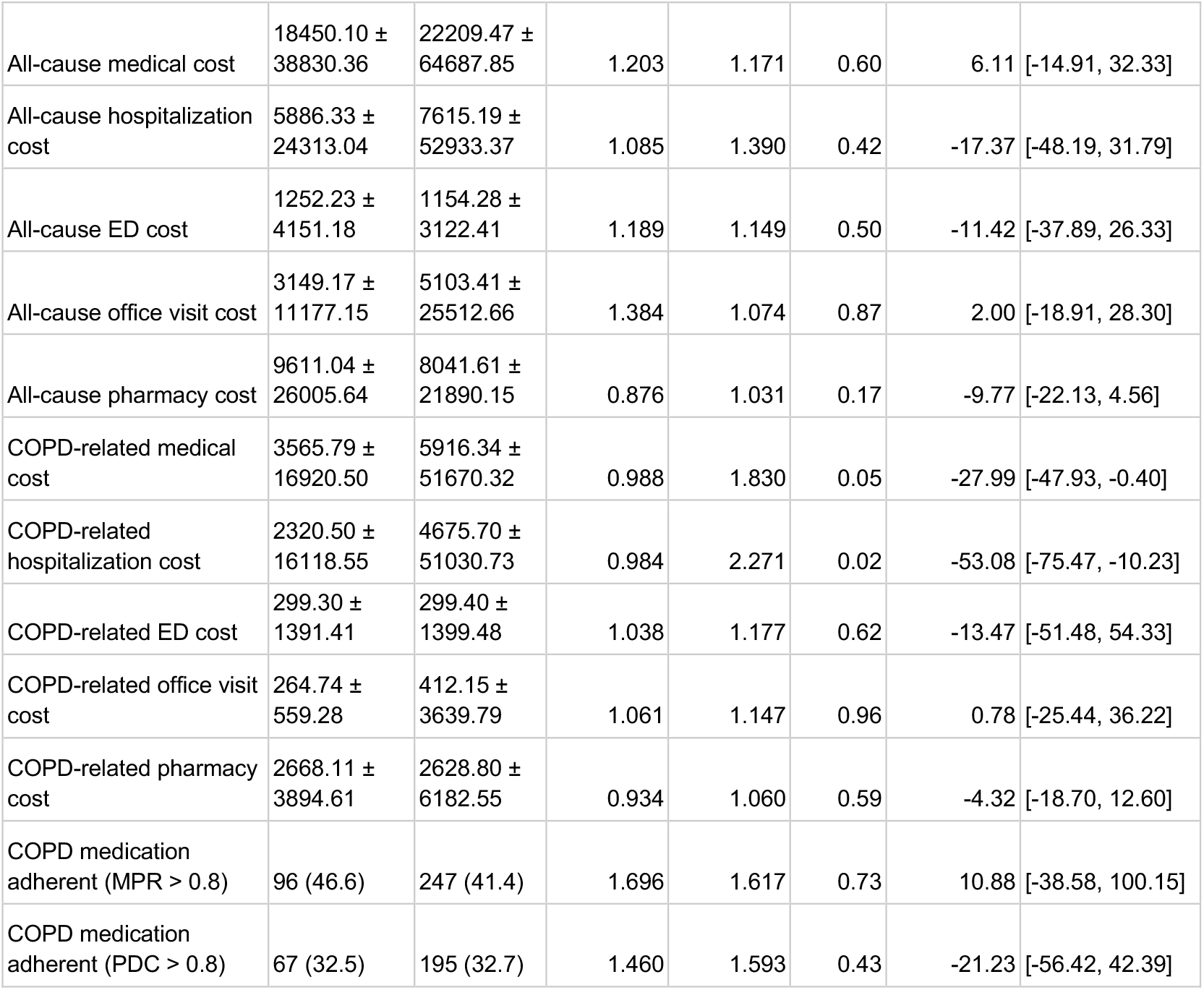
Summary of treatment effects in all-cause and COPD-related utilizations, utilization cost, and pharmacy cost between the intervention and control groups.

**Figure 2.**
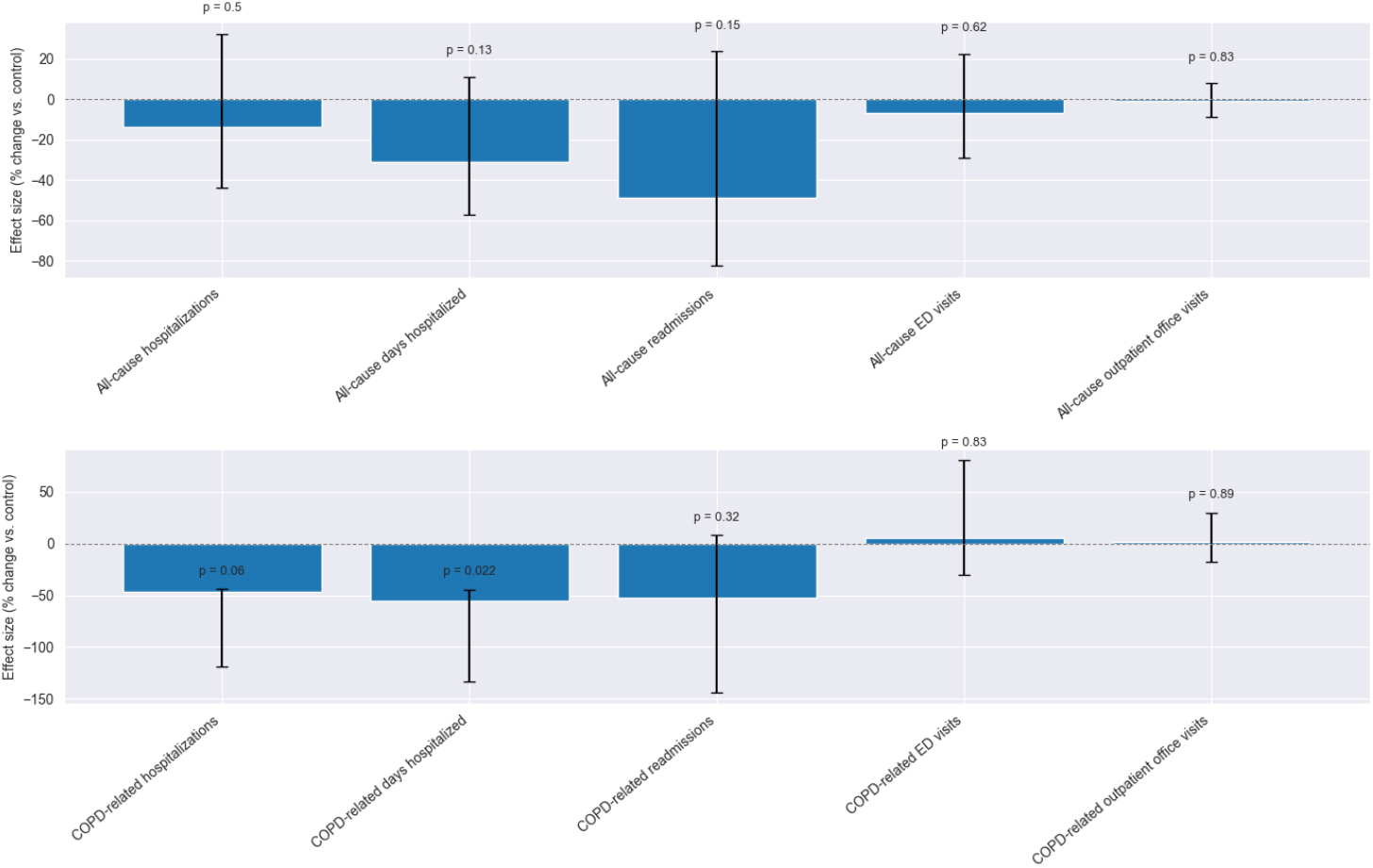
Preliminary results on treatment effects in all-cause and COPD-related utilizations. The effect sizes (height of the bar) and the 95% confidence intervals were obtained using the coefficient estimates from the DiD models for all variables except readmissions. The effect sizes represent the percent change in the annualized post/pre rate ratios for the intervention group compared to the controls in the DID model. A negative value represents a reduction in the rate ratios. For the number of readmissions, effect size and 95% confidence interval was obtained using a Poisson test on the post 6-month period. The effect size for the number of readmissions represents the percentage change in the incidence rate for the intervention group compared to the controls.

**Figure 3.**
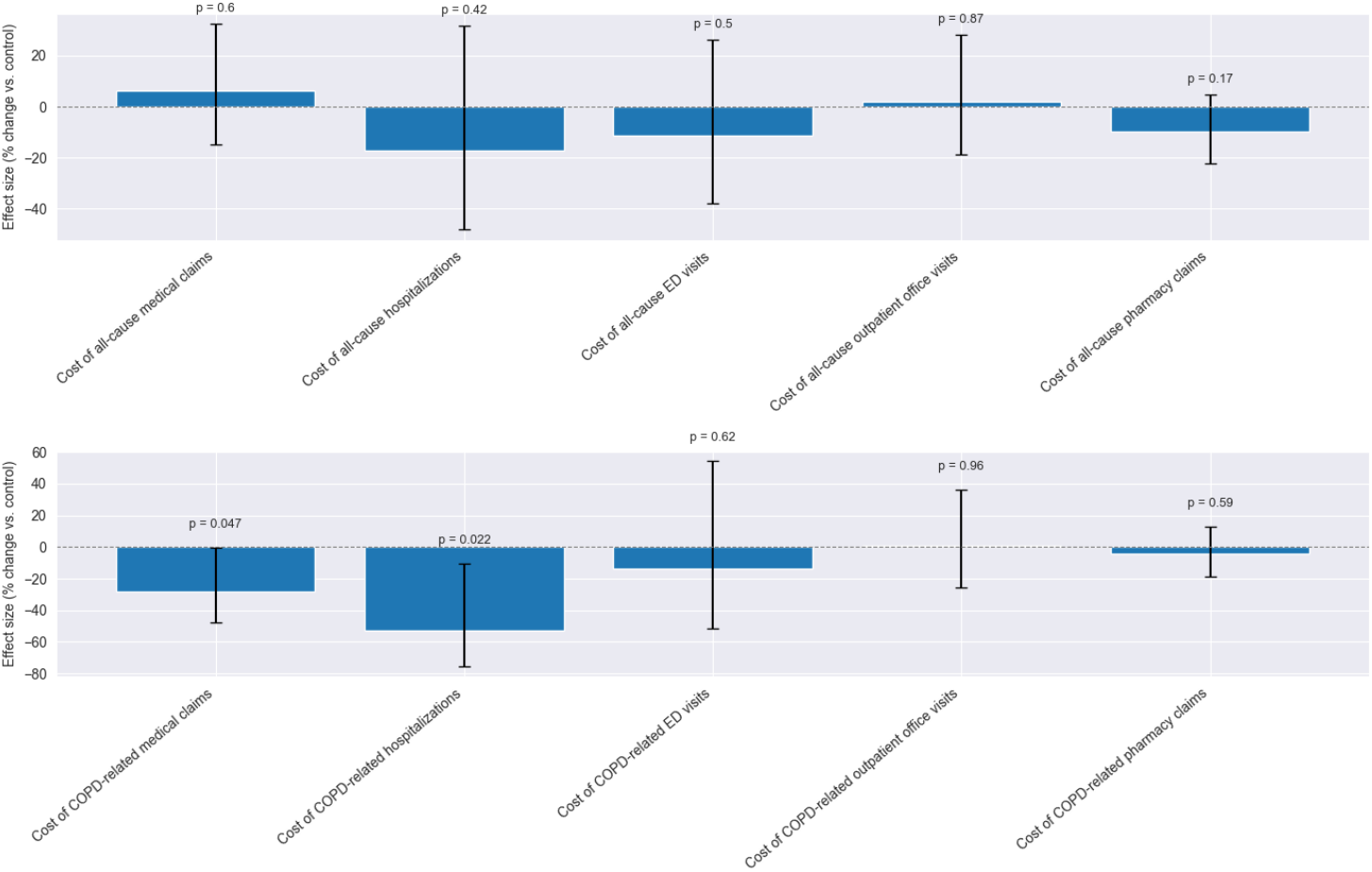
Preliminary results on treatment effects in all-cause and COPD-related utilization cost and pharmacy cost. The effect sizes (height of the bar) and the 95% confidence intervals were obtained using the coefficient estimates from the GLMM with a Tweedie distribution. The effect sizes represent the ratio of cost ratios in the DID model. A negative value represents a reduction in the post/pre cost ratios.

In the sensitivity analyses, similar magnitude of effect sizes and p-value were found using both the bootstrap process described above, and by directly comparing the rate or cost in the 6 months follow-up period between the intervention group and the control group using a GLMM (Table 4). These results reassure that the results are not sensitive to the model assumptions, nor the matched dataset used in the original DID model.

**Table 4.** Comparison of effect sizes and p-values from the DID model, the bootstrapped DID model, and the case-control model for the sensitivity analyses.

| Characteristic | Effect size DID model | P-value DID model | Effect size DID bootstrap | P-value DID bootstrap | Effect size CC model | P-value CC model |
| --- | --- | --- | --- | --- | --- | --- |
| All-cause hospitalizations | -13.79 | 0.50 | -20.45 | 0.35 | -21.33 | 0.21 |
| All-cause average days hospitalized | -30.87 | 0.13 | -41.49 | 0.04 | -31.56 | 0.08 |
| All-cause readmissions | -48.57 | 0.15 | -38.23 | 0.41 | -48.57 | 0.15 |
| All-cause ED visits | -6.63 | 0.62 | 2.15 | 0.78 | 4.11 | 0.75 |
| All-cause outpatient office visits | -0.93 | 0.83 | 0.21 | 0.98 | 0.35 | 0.94 |
| COPD-related hospitalizations | -46.84 | 0.06 | -47.94 | 0.03 | -43.38 | 0.05 |
| COPD-related average days hospitalized | -55.40 | 0.02 | -57.88 | 0.03 | -53.66 | 0.02 |
| COPD-related readmissions | -52.63 | 0.32 | -37.71 | 0.51 | -52.63 | 0.32 |
| COPD-related ED | 5.68 | 0.83 | 11.75 | 0.79 | 8.90 | 0.72 |
| COPD-related outpatient office visit | 1.60 | 0.89 | 9.29 | 0.52 | 1.79 | 0.88 |
| Acute COPD outpatient office visit | 8.53 | 0.71 | 1.11 | 0.88 | -19.65 | 0.30 |
| All-cause medical cost | 6.11 | 0.60 | 4.05 | 0.73 | -9.65 | 0.31 |
| All-cause hospitalization cost | -17.37 | 0.42 | -23.78 | 0.38 | -29.03 | 0.12 |
| All-cause ED cost | -11.42 | 0.50 | 1.50 | 0.95 | 10.22 | 0.55 |
| All-cause outpatient office visit cost | 2.00 | 0.87 | 2.48 | 0.78 | -3.87 | 0.69 |
| All-cause pharmacy cost | -9.77 | 0.17 | -11.23 | 0.23 | 12.91 | 0.24 |
| COPD-related medical cost | -27.99 | 0.05 | -27.43 | 0.06 | -24.36 | 0.12 |
| COPD-related hospitalization cost | -53.08 | 0.02 | -47.26 | 0.14 | -52.42 | 0.03 |
| COPD-related ED cost | -13.47 | 0.62 | 4.25 | 0.91 | 25.37 | 0.42 |
| COPD-related outpatient office visit cost | 0.78 | 0.96 | 19.44 | 0.39 | -15.52 | 0.23 |
| COPD-related pharmacy cost | -4.32 | 0.59 | -0.32 | 0.90 | 10.39 | 0.39 |
| COPD medication adherent (MPR > 0.8) | 10.88 | 0.73 | -1.72 | 0.98 | 26.34 | 0.16 |
| COPD medication adherent (PDC > 0.8) | -21.23 | 0.43 | -36.58 | 0.37 | 0.33 | 0.99 |

Among 840 controls and 280 intervention patients, no patient safety events were detected during the program to date.

The first two columns show the annualized mean +/-SD per patient in the 6-month follow-up period or the number of patients (percentage) for the binary variables (medication adherent). For event counts, this is the incidence rate per patient per year. For cost variables, the mean is the PMPY cost for each category. Ratios are referring to the post/pre rate ratios, cost ratios, or odds ratios (for binary variables). P-value and percent change in ratios was from the exponentiated estimate of the interaction term from the DiD models minus 1 for all variables except number of readmissions. The percent change was the rate ratio estimate from the Poisson test for number of readmissions.

The effect size in the DID model represents the percent change in the annualized post/pre rate(cost) ratios for the intervention group compared to the controls; the effect size in the bootstrap analyses represents the averaged percentage change in the annualized post/pre rate ratio across 200 bootstraps; the effect size in the case-control model represents the percentage change in the annualized rate (cost) in the intervention group compared to the controls. Note that the same model was used for readmission rates in the DID model and the case-control model listed here.

## 6. Discussion

COPD is an important driver of potentially avoidable death, disability, hospitalization, and health care spending and yet access to high quality care is lacking for many Americans. The NuvoAir Home Service provides a commercially viable remote service that is available to patients with COPD from the convenience of their home who otherwise struggle to access medical care. To our knowledge, this is the first large-scale evaluation of a commercially available clinical service to demonstrate the impact of remote physiological monitoring combined with integrated chronic care management and telemedicine. The results show clinically significant and statistically significant improvements in the expected direction, shifting utilization from the inpatient to outpatient setting. Similar results have been observed in other remote COPD management programs.[21,22] Strengths of the study include generalizability: The care-delivery model utilized by the NuvoAir clinical team was designed carefully to optimize effectiveness and convenience for all patients diagnosed with COPD, including those with limited technical and health literacy and including those in rural areas. Smartphone applications and blue-tooth spirometer along with cellular enabled pulse oximetry allows symptom-reporting, monitoring and patientcare to happen anywhere, which improves the adherence to the program.

One major limitation of this study is that COPD diagnoses and severity were not uniformly confirmed by spirometry, therefore some patients without airflow limitation may have been diagnosed with COPD. We could not consistently obtain more comprehensive symptom surveys such as St. George Respiratory Questionnaires from all participants, limiting our ability to measure improvements in quality of life and related measures. Therefore, some exploratory endpoints were not captured as planned. Another limitation of the study is that because the evaluation was not randomized and blinded, patients who were willing to enroll in the program may have more motivation to follow health advice, which could bias results. However, disease-related variables were matched carefully to ensure comparable baseline levels, which should limit possible self-selection bias.

## 7. Conclusion

This report suggests that the NuvoAir Home Service may be effective in reducing COPD-related hospital admissions, average days of hospital stay, readmissions, and related medical costs. While not all endpoints reached statistical significance, promising positive effects are demonstrated with a relatively small number of patients enrolled. These results align with the findings that a well-structured remote care-management program can improve important health outcomes for patients with COPD.

## Data Availability

The data supporting this evaluation will not be publicly available.

## Author Contributions

Eric Harker, Jeffrey Van Wormer, and Dandi Qiao participated in the conception and design of the study, the acquisition of the data, data analysis and/or interpretation, writing and editing of the article.

## Data Sharing

The data supporting this evaluation will not be publicly available.

### Declaration of Interest

Both Eric Harker and Dandi Qiao are employees of NuvoAir Medical Inc, Boston MA. Jeffrey Van Wormer is an employee of Marshfield Clinic Health System, Marshfield WI.

## Abbreviations

PROMISE: NAHS, COPD, CMS, RPM, CCM, ICD10, IRB, GOLD, PSM, GLMM.

## Statement of Funding Support

This evaluation and all related work were funded by the sponsor, NuvoAir Medical Inc.

ClinicalTrials.gov ID NCT05955482.

## Notes

### Clinical Protocols

https://clinicaltrials.gov/study/NCT05955482

### Author Declarations

This study was a quality improvement program evaluation and determined to be exempt by the Marshfield Clinic Health System Institutional Review Board.

## References

1. Saloni Dattani, Fiona Spooner, Hannah Ritchie and Max Roser (2023) - “Causes of Death” Published online at OurWorldinData.org. Accessed Nov 3, 2024. https://ourworldindata.org/causes-of-death

2. Global health estimates: Leading causes of DALYs Disease burden, 2000–2021. Published 2024. Accessed Oct 29, 2024. https://www.who.int/data/gho/data/themes/mortality-and-global-health-estimates/global-health-estimates-leading-causes-of-dalys

3. Mannino D, Siddall J, Small M, et al. Treatment Patterns for Chronic Obstructive Pulmonary Disease (COPD) in the United States: Results From an Observational Cross-Sectional Physician and Patient Survey. International Journal of Chronic Obstructive Pulmonary Disease. 2022;17:749–761. doi:10.2147/COPD.S340794.

4. Anzueto A, Rogers S, Donato B, et al. Treatment Patterns in Patients With Newly Diagnosed COPD in the USA. BMC Pulmonary Medicine. 2024;24(1):395.doi:10.1186/s12890-024-03194-4

5. Gaffney AW, Hawks L, White AC, et al. Health Care Disparities Across the Urban-Rural Divide: A National Study of Individuals With COPD. The Journal of Rural Health : Official Journal of the American Rural Health Association and the National Rural Health Care Association. 2022;38(1): 207–216. doi:10.1111/jrh.12525.

6. Torabipour A, et al. Cost Analysis of Hospitalized Patients with Chronic Obstructive Pulmonary Disease: A State-Level Cross-Sectional Study. Tanaffos. 2016;15(2):75–82.

7. Müllerová H, et al. Association of COPD exacerbations and acute cardiovascular events: a systematic review and meta-analysis. Ther Adv Respir Dis. 2022 Jan-Dec;16:17534666221113647. doi: 10.1177/17534666221113647.

8. Putcha N, Drummond MB, Wise RA, Hansel NN. Comorbidities and Chronic Obstructive Pulmonary Disease: Prevalence, Influence on Outcomes, and Management. Semin Respir Crit Care Med. 2015 Aug;36(4):575–91. doi: 10.1055/s-0035-1556063. Epub 2015 Aug 3. PMID: 26238643; PMCID: PMC5004772.

9. Koff KB, Min S, Diaz DLP, et al. Impact of Proactive Integrated Care on chronic obstructive pulmonary disease. Chronic Obstr Pulm Dis. 2021; 8(1): 100–116. doi: 10.15326/jcopdf.2020.0139

10. Aboumatar H, Naqibuddin M, Chung S, et al. Effect of a hospital-initiated program combining transitional care and long-term self-management support on outcomes of patients hospitalized with chronic obstructive pulmonary disease: a randomized clinical trial. JAMA. 2019;322(14):1371–1380. doi: 10.1001/jama.2019.11982

11. Fan VS, Gaziano JM, Lew R, et al. A comprehensive care management program to prevent chronic obstructive pulmonary disease hospitalizations: a randomized, controlled trial. Ann Intern Med. 2012; 156(10):673–683. doi: 10.7326/0003-4819-156-10-201205150-00003

12. Kerr M, Tarabichi Y, et al. Patterns of care in the management of high-risk COPD in the US (2011-2019): an observational study for the CONQUEST quality improvement program. Lancet Reg Health Am. 2023 Jul 28;24:100546. doi: 10.1016/j.lana.2023.100546. PMID: 37545746; PMCID: PMC10400879.

13. Janjua S, Carter D, Threapleton CJ, Prigmore S, Disler RT. Telehealth Interventions: Remote Monitoring and Consultations for People With Chronic Obstructive Pulmonary Disease (COPD). The Cochrane Database of Systematic Reviews. 2021;7:CD013196. doi:10.1002/14651858.CD013196.pub2.

14. Koh JH, Chong LCY, Koh GCH, Tyagi S. Telemedical Interventions for Chronic Obstructive Pulmonary Disease Management: Umbrella Review. Journal of Medical Internet Research. 2023;25:e33185. doi:10.2196/33185.

15. Watz, H., et al. Spirometric changes during exacerbations of COPD: a post hoc analysis of the WISDOM trial. Respir Res 19, 251 (2018). doi.org/10.1186/s12931-018-0944-3

16. GLOBAL STRATEGY FOR PREVENTION, DIAGNOSIS AND MANAGEMENT OF COPD: 2024 GOLD Report. Published 2024. Accessed Oct 29, 2024. https://goldcopd.org/2024-gold-report/

17. COPD Foundation Educational Materials. https://www.copdfoundation.org/Learn-More/Educational-Materials-Resources/Downloads.aspx

18. Caliendo, M. and Kopeinig, S. (2008), Some practical guidance for the implementation of propensity score matching. Journal of Economic Surveys, 22:31–72. 10.1111/j.1467-6419.2007.00527.x

19. Whaley, C. M., Bollyky, J. B., Lu, W., Painter, S., Schneider, J., Zhao, Z., … Meadows, E. S. (2019). Reduced medical spending associated with increased use of a remote diabetes management program and lower mean blood glucose values. Journal of Medical Economics, 22(9), 869–877. 10.1080/13696998.2019.1609483

20. Wing C, Simon K, Bello-Gomez RA. Designing Difference in Difference Studies: Best Practices for Public Health Policy Research. Annu Rev Public Health. 2018 Apr 1;39:453–469. doi: 10.1146/annurev-publhealth-040617-013507. Epub 2018 Jan 12. PMID: 29328877.

21. Brazeal T, Kaye L, Vuong V, Le J, Peris Z, Barrett MA. Reducing Health Care Resource Utilization in COPD: A Retrospective Matched Control Analysis of a Digital Quality Improvement Program. Chronic Obstr Pulm Dis. 2024 Sep 27;11(5):515–523. doi: 10.15326/jcopdf.2024.0532. PMID: 39242089; PMCID: PMC11548968.

22. Singh M, Hsu ES, Polychronopoulou E, Sharma G, Duarte AG. Structured evaluation and management of patients with COPD in an accredited program. Chronic Obstr Pulm Dis. 2023; 10(3): 297–307. doi: 10.15326/jcopdf.2022.0366

