## Supplemental Tables for "Study Report from the Pragmatic Assessment of the NuvoAir Clinical Service in the Management of Patients with Chronic Obstructive Pulmonary Disease"

Supplemental Files

**Supplemental Tables**

**Supplemental Table 1. Additional summary statistics on cost variables before PSM.**

| **Characteristic** | **Metric** | **Intervention** | **Control** | **P value** |
| --- | --- | --- | --- | --- |
| All-cause medical cost | % with any cost | 100.00% | 100.00% | 1 |
| All-cause medical cost | Median (Q1-Q3) if >0 | 6,499.99  (3,077.97–17,685.50) | 5,201.37  (1,961.57–17,738.21) | 0.00109 |
| All-cause hospitalization cost | % with any cost | 21.80% | 22.30% | 0.899 |
| All-cause hospitalization cost | Median (Q1-Q3) if >0 | 19,331.30  (13,610.88–33,465.90) | 16,755.28  (10,178.47–29,875.97) | 0.128 |
| All-cause ED cost | % with any cost | 45.00% | 38.40% | 0.0273 |
| All-cause ED cost | Median (Q1-Q3) if >0 | 1,353.02  (688.76–3,215.81) | 1,210.87  (606.35–3,005.26) | 0.408 |
| All-cause office visit cost | % with any cost | 96.80% | 96.50% | 0.941 |
| All-cause office visit cost | Median (Q1-Q3) if >0 | 1,741.88  (982.09–2,626.69) | 1,224.34  (637.69–2,240.39) | 2.22E-08 |
| All-cause pharmacy cost | % with any cost | 87.50% | 82.50% | 0.0345 |
| All-cause pharmacy cost | Median (Q1-Q3) if >0 | 3,828.17  (484.83–11,483.39) | 2,068.47  (314.18–7,146.00) | 1.22E-05 |
| COPD-related medical cost | % with any cost | 69.60% | 54.60% | 6.61E-07 |
| COPD-related medical cost | Median (Q1-Q3) if >0 | 975.47  (375.00–3,391.38) | 713.65  (319.82–4,269.56) | 0.0262 |
| COPD-related hospitalization cost | % with any cost | 12.50% | 11.60% | 0.688 |
| COPD-related hospitalization cost | Median (Q1-Q3) if >0 | 17,208.82  (11,024.77–21,065.44) | 14,661.65  (9,735.38–23,977.61) | 0.761 |
| COPD-related ED cost | % with any cost | 17.10% | 9.90% | 7.24E-05 |
| COPD-related ED cost | Median (Q1-Q3) if >0 | 1,156.05  (837.72–1,933.39) | 1,132.70  (738.43–2,454.11) | 0.941 |
| COPD-related office visit cost | % with any cost | 49.30% | 34.50% | 2.91E-07 |
| COPD-related office visit cost | Median (Q1-Q3) if >0 | 421.13  (207.22–663.18) | 317.89  (184.86–552.63) | 0.0122 |
| COPD-related pharmacy cost | % with any cost | 71.10% | 58.90% | 4.49E-05 |
| COPD-related pharmacy cost | Median (Q1-Q3) if >0 | 2,525.84  (303.50–6,750.39) | 1,159.85  (106.55–4,351.93) | 4.16E-06 |

**Supplemental Table 2. Additional summary statistics on cost variables after PSM.**

| **Characteristic** | **Metric** | **Intervention** | **Control** | **P value** |
| --- | --- | --- | --- | --- |
| All-cause medical cost | % with any cost | 100.00% | 100.00% | 1 |
| All-cause medical cost | Median (Q1-Q3) if >0 in $ | 6,499.99  (3,077.97–17,685.50) | 8,150.57 (2,759.47–21,927.89) | 0.682 |
| All-cause hospitalization cost | % with any cost | 21.80% | 23.90% | 0.514 |
| All-cause hospitalization cost | Median (Q1-Q3) if >0 in $ | 19,331.30  (13,610.88–33,465.90) | 16,713.86 (10,540.67–26,925.57) | 0.134 |
| All-cause ED cost | % with any cost | 45.00% | 40.10% | 0.172 |
| All-cause ED cost | Median (Q1-Q3) if >0 in $ | 1,353.02  (688.76–3,215.81) | 1,275.53 (638.70–3,104.15) | 0.81 |
| All-cause office visit cost | % with any cost | 96.80% | 98.50% | 0.136 |
| All-cause office visit cost | Median (Q1-Q3) if >0 in $ | 1,741.88  (982.09–2,626.69) | 1,565.26 (874.11–2,896.15) | 0.481 |
| All-cause pharmacy cost | % with any cost | 87.50% | 87.00% | 0.918 |
| All-cause pharmacy cost | Median (Q1-Q3) if >0 in $ | 3,828.17  (484.83–11,483.39) | 3,547.43 (595.12–8,932.27) | 0.31 |
| COPD-related medical cost | % with any cost | 69.60% | 68.30% | 0.738 |
| COPD-related medical cost | Median (Q1-Q3) if >0 in $ | 975.47  (375.00–3,391.38) | 899.19 (342.94–3,273.04) | 0.129 |
| COPD-related hospitalization cost | % with any cost | 12.50% | 11.40% | 0.707 |
| COPD-related hospitalization cost | Median (Q1-Q3) if >0 in $ | 17,208.82 (11,024.77–21,065.44) | 12,942.70 (9,067.54–19,461.81) | 0.249 |
| COPD-related ED cost | % with any cost | 17.10% | 15.10% | 0.476 |
| COPD-related ED cost | Median (Q1-Q3) if >0 in $ | 1,156.05  (837.72–1,933.39) | 1,180.31 (727.73–2,089.82) | 0.983 |
| COPD-related office visit cost | % with any cost | 49.30% | 49.20% | 1 |
| COPD-related office visit cost | Median (Q1-Q3) if >0 in $ | 421.13  (207.22–663.18) | 383.98 (210.17–689.90) | 0.858 |
| COPD-related pharmacy cost | % with any cost | 71.10% | 71.50% | 0.939 |
| COPD-related pharmacy cost | Median (Q1-Q3) if >0 in $ | 2,525.84  (303.50–6,750.39) | 1,827.08 (284.70–5,219.90) | 0.0713 |
